# Motivational Strategies for Stroke Rehabilitation: A Descriptive Cross-sectional Study

**DOI:** 10.1101/19011023

**Authors:** Kazuaki Oyake, Makoto Suzuki, Yokei Otaka, Satoshi Tanaka

## Abstract

**Background and Purpose:** The addition of motivational strategies to a rehabilitation program is thought to enhance patient adherence and improve outcomes. However, little is known about how rehabilitation professionals motivate stroke patients during rehabilitation. The primary objective of this study was to provide a comprehensive and quantitative list of motivational strategies for stroke rehabilitation. In addition, we aimed to examine (1) whether professionals with more clinical experience used a higher number of motivational strategies, (2) the purpose for using each strategy, and (3) the information considered when choosing strategies.

**Methods:** This descriptive, cross-sectional study was conducted using a web survey with a random sample of 407 rehabilitation professionals including physicians, nurses, physical therapists, occupational therapists, and speech-language-hearing therapists.

**Results:** We received data for 362 participants. Fifteen strategies were found to be used by more than 75% of the respondents reported using to motivate their patients. Almost all of the respondents reported that they actively listen to and praise their patients to increase patient adherence to rehabilitation programs. Respondents with more clinical experience tended to use a higher number of motivational strategies (rho = 0.208, p < 0.001). For 11 of the 15 strategies selected by more than 75% of respondents, the highest percentage of respondents reported that they used the strategies to make rehabilitation worthwhile for their patients. The majority of respondents reported that they decide which motivational strategy to use by considering comprehensive information regarding the patient health condition, environmental factors, and personal factors.

**Conclusions:** The comprehensive list of motivational strategies obtained may be useful for increasing patient adherence to rehabilitation, especially for professionals with less clinical experience. Furthermore, our findings regarding the purpose for using each strategy and the information used to choose strategies might help rehabilitation professionals to utilize the motivational strategy list.

## Introduction

Studies on stroke rehabilitation have recommended the use of intensive and repetitive task-specific practice, as well as aerobic exercise.^1^ Given that the patient’s own efforts are necessary to sustain these practices and exercises, patient motivation is frequently used as a determinant of rehabilitation outcome.^2^ High adherence to a rehabilitation program is thought to be indicative of motivation,^2, 3^ and a lack of motivation is one of the perceived barriers to physical activity and exercise training after stroke.^4-7^ Therefore, the addition of motivational strategies to rehabilitation programs may effectively enhance patient adherence, producing better outcomes.^8^

Motivational strategies such as feedback,^9, 10^ counseling,^11^ and information provision^12^ have positive effects on recovery after stroke. An international randomized clinical trial found that praise and positive feedback were effective for improving walking speed during inpatient rehabilitation.^9^ Feedback using virtual reality has been found to be beneficial in improving motivation, upper limb function, and activities of daily living.^10,13^ Counseling and information provision have a positive effect of mood.^11, 12^ However, few reports have comprehensively investigated strategies used by medical professionals to motivate patients undergoing stroke rehabilitation.

Maclean et al. ^3^ conducted a semi-structured interview of medical professionals to determine how they increase patient motivation with respect to stroke rehabilitation. The researchers reported that setting rehabilitation goals, providing information about rehabilitation, and accessing and using the patient’s cultural norms appeared to have a positive effect on motivation.^3^ However, the generalizability of these findings was limited due to the nature of qualitative research.^14^

As opposed to semi-structured interviews, the findings from quantitative surveys are generalizable to a larger population.^14^ Therefore, the primary objective of this study was to provide a comprehensive list of motivational strategies that medical professionals use for stroke rehabilitation based on quantitative survey data. A list of motivational strategies is likely to be useful in increasing patient adherence to rehabilitation. We hypothesized that motivational skills could be acquired through clinical experience. In addition, understanding the purpose of each motivational strategy and the information that is evaluated when choosing strategies may contribute to effective utilization of the list. Thus, our secondary objectives were to examine (1) whether rehabilitation professionals with greater clinical experience used more motivational strategies, (2) the purpose for using each strategy, and (3) the information considered when choosing motivational strategies.

## Methods

### Data availability statement

The data that support the findings of this study are available from the corresponding author on reasonable request.

### Study design

This study had a descriptive cross-sectional design. We used a random sampling web-based survey to obtain quantitative results from the participant perspective. The study protocol was approved by the appropriate ethics committee at the Hamamatsu University School of Medicine (approval number: 18-136). Informed consent was obtained from all participants.

### Participants

Eligible participants were rehabilitation professionals including physicians, nurses, physical therapists, occupational therapists, speech-language-hearing therapists, or clinical psychologists currently working in rehabilitation. Participants were recruited with the cooperation of the 33rd conference of the Comprehensive Rehabilitation Ward Association, the 48th Annual Meeting of the Nagano Physical Therapy Association, and other rehabilitation-related associations. To recruit participants, we used leaflets and posters containing a brief description of the survey and a hyperlink to the survey, which was conducted online. The first page of the survey informed participants about the total number of questions, the approximate response time, and the aim of the study. Participants were asked to report their professional category and years of experience in stroke rehabilitation. Those who met the eligible criteria proceeded to the next page.

### Survey instrument

We consulted the guidelines and followed a checklist to prepare the survey.^15, 16^ We chose to use a voluntarily accessed survey developed by using the Google Forms tool (Google LLC, Mountain View, CA, USA). Among the individuals who completed the survey, 50 were selected via a draw to receive an honorarium of 1,000 yen (approximately US $9.00).

The survey items regarding motivational strategies were developed based on the clinical experience of the authors, the findings of related literature,^2-5, 7, 9, 11, 12, 17-20^ and data obtained from semi-structured interviews with five professionals about motivational strategies for stroke rehabilitation.^21^ The experts in stroke rehabilitation reviewed the items to ensure the content validity. We carried out a pilot test with a group of 10 rehabilitation professionals to determine whether the respondents understood the questions and instructions, and whether the meaning of the questions was the same for all respondents.^16^ The survey took an average of 15 min to complete and the qualitative feedback from the semi-structured interviews with the 10 respondents resulted in some minor grammatical changes. Consequently, we prepared a list of 22 motivational strategies (Table 1). In the first section, respondents were shown the list of motivational strategies and asked if they used each strategy in clinical practice. Respondents were also asked to respond to an open-ended question when they were invited to propose additional motivational strategies that were not included in the list.

**Table 1.** List of motivational strategies

| Motivational strategy | Representation in the manuscript |
| --- | --- |
| Active listening | Active listening |
| Allowing the patient to use a newly acquired skill | Allowing the patient to use a newly acquired skill |
| Applying patient preferences to practice and exercise tasks | Application of patient's preferences |
| Control of task difficulty | Control of task difficulty |
| Explaining the necessity of a practice | Explaining the necessity of a practice |
| Goal-oriented practice | Goal-oriented practice |
| Goal setting | Goal setting |
| Group rehabilitation | Group rehabilitation |
| Engaging in enjoyable communication with the patient | Enjoyable communication |
| Praise | Praise |
| Proposing conditions for exchange (e.g. promising to undertake the patient's favorite practice after achieving his/her least favorite practice) | Proposing conditions for exchange |
| Providing a suitable rehabilitation environment | Providing a suitable rehabilitation environment |
| Providing exercise and practice with game properties | Practice with game properties |
| Providing medical information | Providing medical information |
| Providing opportunities for the patient to identify possible treatments | Providing opportunities to identify possible treatments |
| Providing practice tasks relating to the patient's experience and lifestyle | Practice related to patient's experience |
| Providing variations of rehabilitation programs to sustain interest | Providing variations of the program |
| Recommending the presence of a family member during rehabilitation | Family member present |
| Respect for self-determination | Respect for self-determination |
| Sharing the criteria for evaluation | Sharing the criteria for evaluation |
| Specifying the amount of practice and exercise that will be required | Specifying the amount of practice required |
| Using tools such as a diary or graph that enables the patient to track his/her progress | Using progress-confirming tools |

Motivational strategies are arranged in alphabetical order.

In the second section, for each motivational strategy that a respondent used, they were asked to state their aim of using the strategy from the following four purposes: (1) to increase the patient’s interest in rehabilitation; (2) to make rehabilitation worthwhile for the patient; (3) to help the patient gain confidence in performing a rehabilitation task; and (4) to increase patient satisfaction with rehabilitation. These purposes were based on the four sub-components of motivation proposed by the Attention, Relevance, Confidence, and Satisfaction (ARCS) model.^22, 23^ The ARCS model is a problem solving approach of designing motivational aspects of learning environments with the goal of stimulating and sustaining students’ motivation to learn.^22^ In the third section, respondents were shown 11 items related to the health conditions of the patients, environmental factors, and personal factors (Table I in the online-only Data Supplement).^24^ They were also asked to select all of the items that they considered when deciding which motivational strategy to use. Finally, respondents were asked to report their sex, primary affiliation, and the phase of stroke recovery of the patients they worked with.

### Sample size

We used the Cochran’s sample size formula to calculate the sample size. We set an alpha level a priori at 0.05, an acceptable error level of 5%, and a confidence interval of 95%.^25^ Consequently, a minimum of 384 participants were required. Assuming that approximately 5% of the participants were excluded, we aimed to recruit a total of 400 participants.

### Statistical analysis

We used descriptive statistics to characterize the study sample and summarize their responses. The normality of distribution was tested using the Shapiro-Wilk test. We used Pearson’s product-moment correlation coefficient or Spearman’s rank correlation coefficient to test whether respondents with more experience used more motivational strategies. For the motivational strategies that were used by at least 75% of respondents^26^, we conducted a hierarchical cluster analysis using the Ward’s method with a squared Euclidean distance to group them according to the purpose of use. Statistical analyses were performed using the Statistical Package for the Social Sciences software version 25.0 (International Business Machines Corp., NY, USA). Any p values < 0.05 were considered statistically significant.

### Reporting

This study was reported according to the recommended best practices and guidelines in the literature for the reporting of survey research.^16, 27^

## Results

The survey was conducted from February to July 2019. Among the 407 participants who completed the survey, 45 respondents were excluded because they did not meet the eligibility criteria, refused to participate, or had missing data. Consequently, 362 respondents completed the survey. The flow of participants is shown in Figure 1. The majority of the respondents were physical therapists (51.1%), had less than 5 years of clinical experience (36.2%), were female (53.0%), were employed in a hospital (91.2%), and worked with patients with subacute stroke (71.3%). The distributions of respondents according to professional category and years in rehabilitation practice are presented in Figure 2. No clinical psychologist participated in the survey.

**Figure 1.**
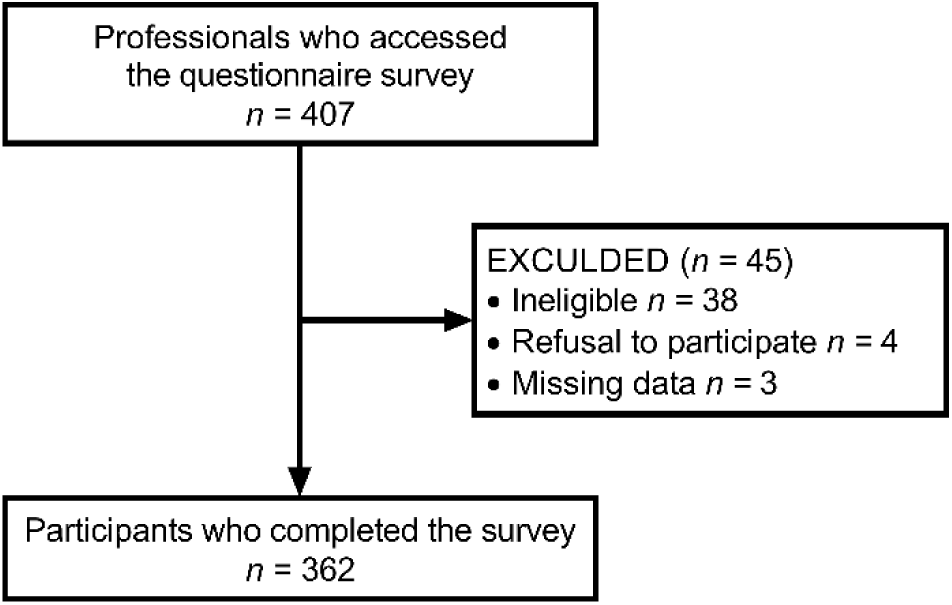
Flow diagram of the participant selection process.

**Figure 2.**
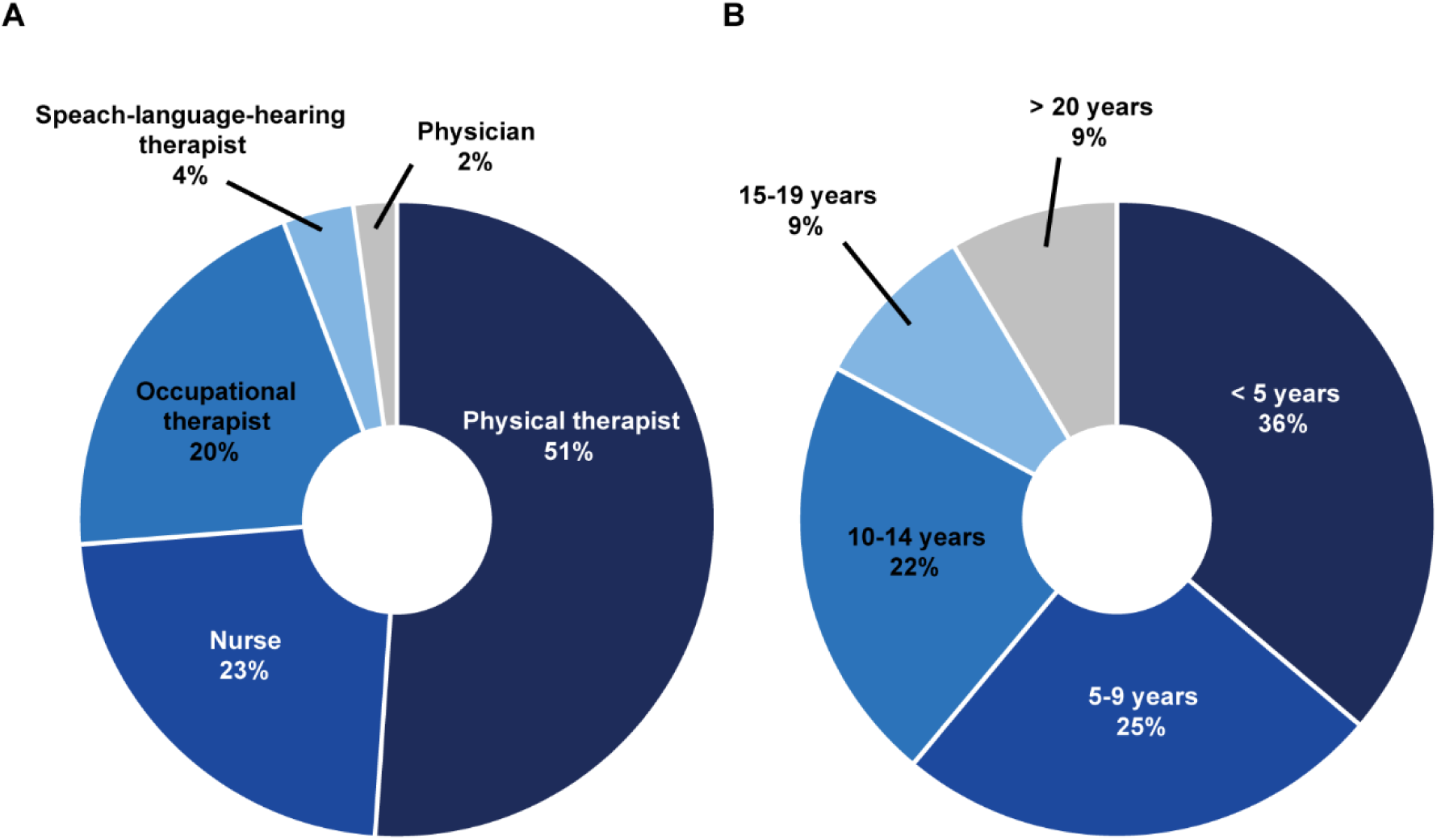
Distribution of respondents according to professional category (A) and years of rehabilitation practice (B).

### Which strategies do rehabilitation professionals use to motivate their patients?

The percentages of respondents who used each of the presented motivational strategies are shown in Figure 3. The majority of the respondents (98.3%) selected “active listening” and “praise”. “Enjoyable communication”, “providing a suitable rehabilitation environment”, “goal setting”, “explaining the necessity of a practice”, and “respect for self-determination” were also selected by more than 90% of the respondents (95.3%– 91.7%). The majority of the respondents reported that they used “control of task difficulty”, “family member present”, “goal-oriented practice”, “providing medical information”, “application of patient preferences”, “practice related to patient’s experience”, “providing opportunities to identify possible treatments”, and “specifying the amount of practice required” (89.2%–76.0%). Thus, 15 out of the 22 presented strategies were selected by more than 75% of the respondents. Between 69.3% and 66.9% of the respondents reported that they used “allowing the patient to use a newly acquired skill”, “sharing the criteria for evaluation”, and “providing variations of the program”. Less than half of the respondents selected “practice with game properties”, “proposing conditions for exchange”, “using progress-confirming tools”, and “group rehabilitation” (49.2%–34.5%). The results by professional category are shown in Figure I in the online-only Data Supplement. The numbers of motivational strategies selected by at least 75% of the respondents who were nurses, physical therapists, and occupational therapists were 10, 15, and 18, respectively. There were four additional motivational strategies proposed by the respondents (Table II in the online-only Data Supplement). Respondents with more clinical experience tended to use more motivational strategies (rho = 0.208, 95% confidence interval = 0.105, 0.308, p < 0.001) (Figure II in the online-only Data Supplement).

**Figure 3.**
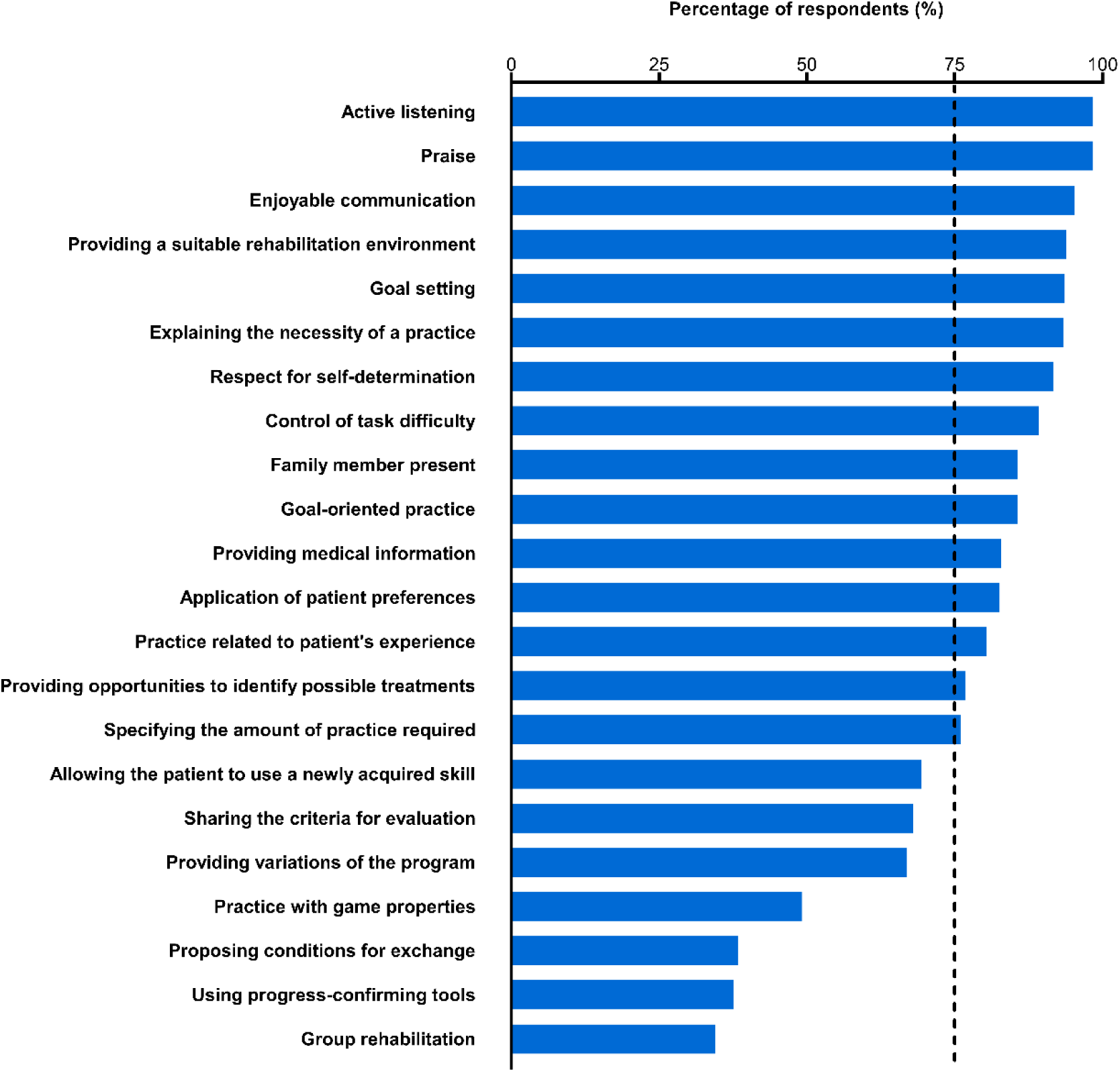
The percentage of respondents who reported that they used each presented motivational strategy during stroke rehabilitation. The vertical dashed line represents 75% of the respondents. The motivational strategies are arranged in descending order by the percentage of respondents who stated that they used each strategy.

### For what purpose do rehabilitation professionals use each motivational strategy?

For 11 of the 15 motivational strategies that were used by at least 75% of the respondents, the highest percentage of respondents reported that they used the strategies to make rehabilitation worthwhile for their patients (47.6%–31.6%) (Figure 4A). For “control of task difficulty” and “praise”, the most common purpose for use was to help the patient gain confidence in performing a rehabilitation task (45.4% and 35.5%, respectively). The largest proportion of respondents who used “enjoyable communication” and “application of patient preferences” reported that they used these strategies to increase the patient’s interest in rehabilitation (49.3% and 39.4%, respectively).

**Figure 4.**
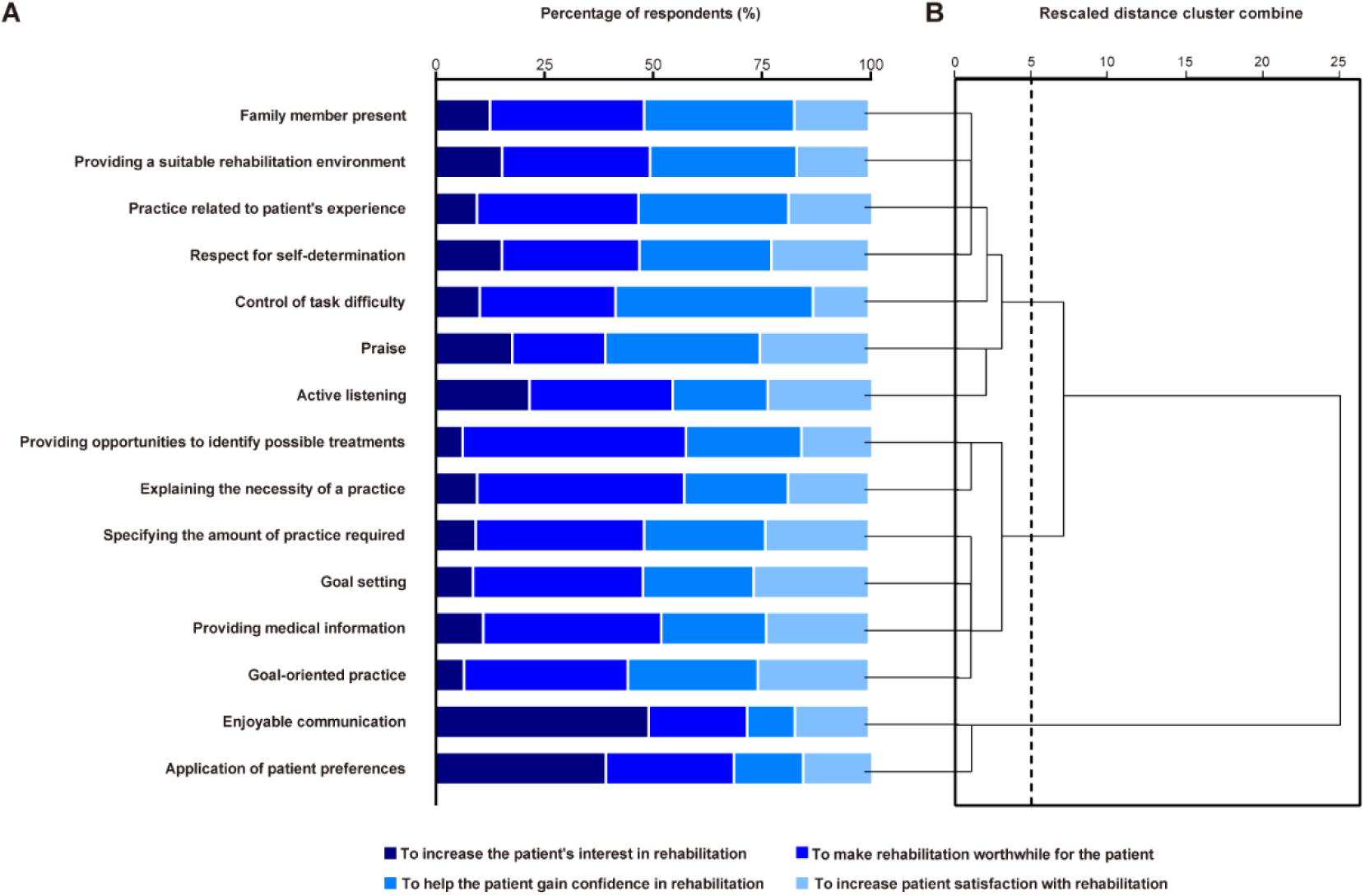
The purpose of using each motivational strategy used by at least 75% of the respondents (A). The order corresponds to Figure 4B. A dendrogram showing the three distinct clusters identified based on hierarchical cluster analysis via Ward’s method (B). The vertical dashed line indicates the optimal number of groups.

The hierarchical cluster analysis produced three motivational strategy clusters (Figure 4B). The first cluster included seven motivational strategies, such as “Family member present” and “providing a suitable rehabilitation environment”. For six of these seven strategies, more than 30% of the respondents reported that they used the strategies to help the patients gain confidence in performing a rehabilitation task. The second cluster included six motivational strategies, such as “providing opportunities to identify possible treatments” and “explaining the necessity of a practice”. Most of the respondents used these to make rehabilitation worthwhile for their patient. The third cluster comprised two strategies that the largest percentage of respondents reported to be using to increase the patient’s interest in rehabilitation.

### What information is considered when rehabilitation professionals choose motivational strategies?

Each of the presented 11 items was selected by more than 75% of respondents as the information that they considered when deciding which motivational strategy to use (Figure 5). Almost all of the respondents selected “patient’s reaction to a presented motivational strategy” and “personality” (97.5%), whereas the lowest percentage of respondents selected “diagnosis” (75.4%). Furthermore, approximately half of the respondents selected all of the 11 presented items (49.1%).

**Figure 5.**
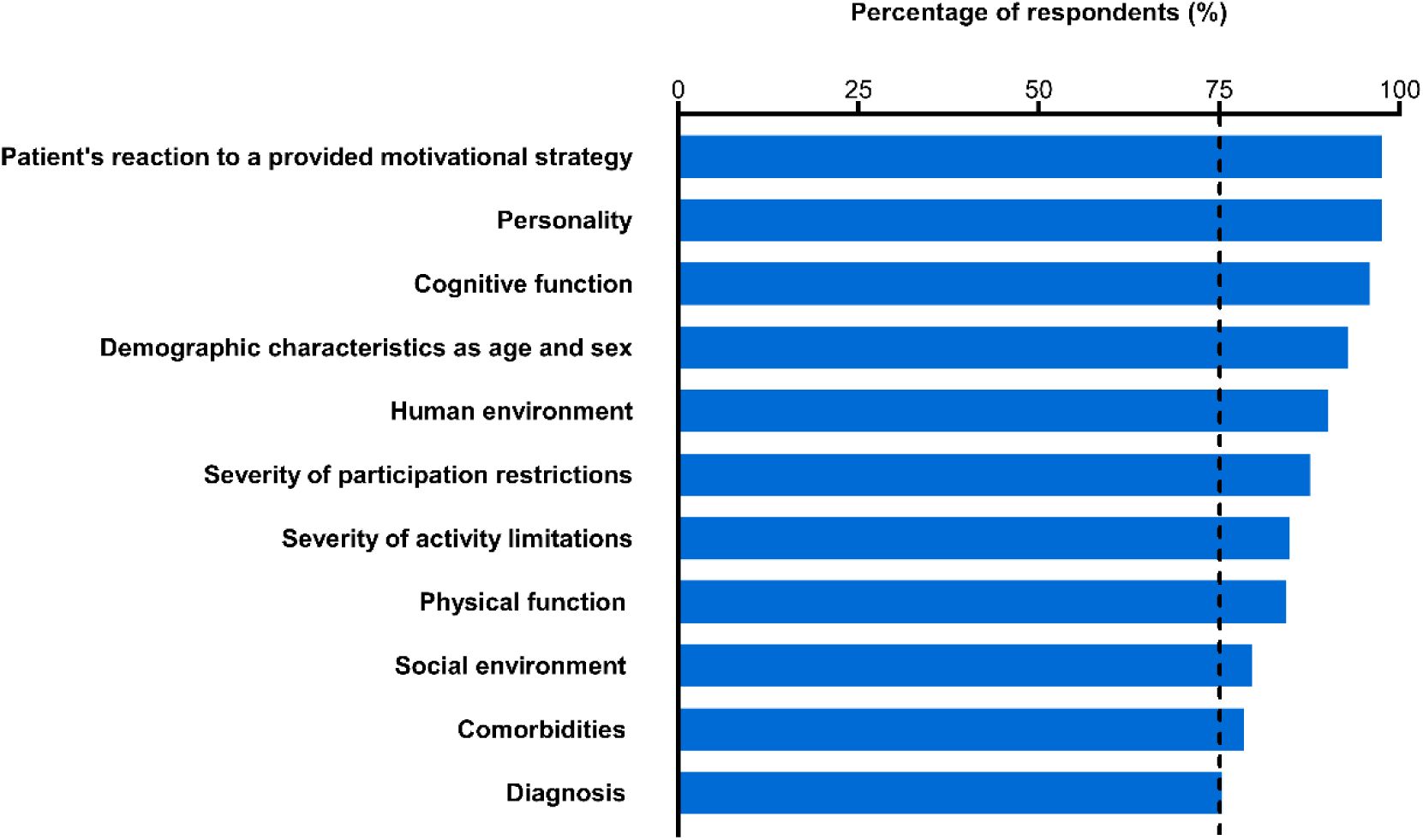
The percentage of respondents who reported to consider each type of information when deciding which motivational strategy to use. Outcomes are arranged in descending order by the percentage of respondents.

## Discussion

In the present study, we generated a comprehensive list of motivational strategies for stroke rehabilitation based on quantitative survey data. We identified 15 motivational strategies used by more than 75% of the respondents. Our results support our hypothesis that rehabilitation professionals acquire skills to motivate stroke patients via clinical experience. Furthermore, our survey generated data regarding the purpose of using each motivational strategy and the information that professionals consider when they choose the motivational strategies. To our knowledge, the present study is the first survey-based research to comprehensively investigate the strategies used to motivate patients with stroke to rehabilitation. We prepared the survey in accordance with the existing guidelines,^15, 16^ and conducted pretesting procedures such as expert reviews and pilot testing to establish content and response process validity.^27^ In addition, we utilized random sampling for data collection. We expected that these procedures would enhance the reliability and generalizability of our findings.

### Which strategies do rehabilitation professionals use to motivate their patients?

Active listening is one of the core communication skills in counseling.^11, 29^ A systematic review reported that counseling has a positive effect on mood in stroke patients.^11, 12^ Praise can induce positive mood and increase motivation to perform a motor skill, resulting in improved performance.^30-32^ Moreover, daily feedback with praise has been reported to be effective in improving walking speed in stroke inpatients.^9^ Almost all of the respondents in our study reported that they actively listen to and praise their stroke patients to increase patient motivation regarding rehabilitation.

Our results suggest that the majority of the surveyed rehabilitation professionals recognized that the communication skills of the medical professional, rehabilitation environment, setting of goals, provision of information, and the presence of family members during practice can affect patient motivation regarding rehabilitation. These findings are consistent with those of Maclean et al..^3^ Communication training programs for clinicians have been shown to increase patient satisfaction, levels of motivation for goal setting and action, and health-related quality of life.^33^ Sonoda et al.^34^ found the providing a specific rehabilitation environment for stroke inpatients increased the amount of physical activity that they engaged in during activities of daily living. Furthermore, providing information about rehabilitation and insuring that a family member is present during rehabilitation may be effective in improving a patient’s mood and encouraging them to be more active.^12, 35^

Respect for self-determination, control of task difficulty, and provision of a goal-oriented practice were motivational strategies that were selected by more than 75% of the respondents. Self-determination contributes to the facilitation and maintenance of intrinsic motivation.^36^ The difficulty of a practice task is thought to have an important effect on the effectiveness of rehabilitation, as inappropriate levels of difficulty can bore or frustrate the patient.^37^ Moreover, gradual increases in task difficulty and goal-oriented practice have been recommended to facilitate functional motor recovery during stroke rehabilitation.^1^ Therefore, through clinical practice, professionals may have observed the effectiveness of these motivational strategies for increasing patient adherence to rehabilitation.

We found the following four strategies that were reportedly used by less than half of the respondents to motivate their patients: providing practice with game properties, proposing conditions for exchange, using progress-confirming tools, and providing group rehabilitation. It remains unclear why these particular motivational strategies were not used. Environmental and time constraints, professional lack of confidence regarding the practice, and lack of perceived effectiveness may prevent rehabilitation professionals from using specific motivational strategies.^28^ In addition, some respondents may not have selected these strategies because they did not understand the accompanying motivational effects, even if they did use these strategies in clinical practice.

### For what purpose do rehabilitation professionals use each motivational strategy?

In clinical settings, rehabilitation professionals may be required to select strategies according to the cause of a patients’ lack of motivation. Therefore, we examined the reasons for using each motivational strategy. For 11 of the 15 strategies used by the majority of respondents, the highest percentage of respondents reported that they used the strategy to make rehabilitation worthwhile for their patient. Lack of knowledge about the potential benefits of training may decrease patient adherence to rehabilitation.^19^ Therefore, professionals may emphasize patient comprehension regarding the benefits and significance of practices to motivate patients regarding rehabilitation.

The hierarchical cluster analysis revealed three groups of motivational strategies. Strategies that center on the value of rehabilitation for patients, such as explaining the necessity of a practice and exercise, are expected to motivate patients with poor understanding regarding the benefits of rehabilitation. For patients with low confidence regarding their practice tasks, strategies focused on increasing patient confidence, such as control of task difficulty and praise, may be effective for increasing patient motivation. Moreover, engaging in enjoyable conversation and applying patient preferences to practice tasks are likely to increase patient interest and prevent the patient from getting bored during rehabilitation. There were no strategies that were used by the majority of respondents to increase patient satisfaction with rehabilitation. However, more than a quarter of the respondents reported that they use goal setting, goal-oriented practice, and praise to increase patient satisfaction with rehabilitation. These strategies may be effective in motivating patients who are not satisfied with rehabilitation. Thus, our findings may help rehabilitation professionals to choose strategies from the list according to the cause of a patients’ lack of motivation.

### What information is considered when rehabilitation professionals choose motivational strategies?

Maclean et al.^3^ reported that compared with younger patients, older unmotivated stroke patients did not respond as well to encouragement. Thus, rehabilitation professionals may benefit from choosing motivational strategies according to the characteristics of each patient. Our results suggest that the majority of rehabilitation professionals choose motivational strategies based on comprehensive data regarding a patient’s health condition, environmental factors, and personal factors. Patient personality type and responses to motivational strategies suggested by the professional seem to be regarded as especially essential information. These findings may assist rehabilitation professionals in deciding which motivational strategy to use and contribute to the effective utilization of the motivational strategy list.

### Limitations

This study had several limitations. First, there may have been a perceived lack of differentiation between the motivational strategies. As our list of motivational strategies comprised specific examples about motivating a patient, some of the presented motivational strategies may have overlapped with one another,^8^ potentially confusing the respondents. However, we designed the motivational strategy list to show specific motivational strategies so that it would be easy to use in a clinical practice. Second, the responses obtained from the survey may not reflect the actual practice of the professional, and may have been affected by inaccurate recall or instead reflect the beliefs or desires of the respondents.^38^ However, we believe that our web-based survey carried a low participant burden and enabled more complete population coverage for sampling.^38, 39^ Finally, all of the participants were recruited from Japan. Whether our findings are generalizable to rehabilitation professionals outside of Japan remains unclear, although the present results are consistent with the previous qualitative^9, 28^ and experimental reports^9, 11, 12, 29-37^ from Western countries. An inter-national survey of motivational strategies for stroke rehabilitation would improve the external validity of our findings.

In summary, we generated a comprehensive and qualitative list of motivational strategies used for stroke rehabilitation, and found that our 15 motivational strategies were used by the majority of rehabilitation professionals. In addition, we obtained data regarding the purpose for using each motivational strategy and the information considered when choosing motivational strategies. These findings may enhance the effective utilization of the motivational strategy list in stroke rehabilitation.

## Acknowledgments

The authors thank Taiki Yoshida, Aiko Akiyama, and Kunitsugu Kondo at Tokyo Bay Rehabilitation Hospital, Keisuke Tani at Hamamatsu University School of Medicine, and Kimito Momose at Shinshu University for their help and support.

